# Bivalent COVID-19 booster vaccines induce cross-reactive but not BA.5-specific antibodies in polyclonal serum

**DOI:** 10.1101/2023.04.12.23288362

**Authors:** Juan Manuel Carreño, Gagandeep Singh, Anass Abbad, Temima Yellin, Komal Srivastava, Charles Gleason, PVI study group, Harm van Bakel, Viviana Simon, Florian Krammer

**Affiliations:** Icahn School of Medicine at Mount Sinai, New York, NY, USA

## Abstract

The question if the bivalent mRNA COVID-19 booster vaccination, containing wild type and BA.5 spike, provides enhanced benefits against BA.5 and similar Omicron subvariants has been widely debated. One concern was an ‘original antigenic sin’-like effect which may redirect immune responses to the bivalent vaccine towards the wild type spike and may block *de novo* generation of BA.5 specific antibodies. Here, we characterized the response to the bivalent vaccine and we performed antibody depletion experiments. Interestingly, when we depleted serum of all antibodies to wild type RBD, we also removed all reactivity to BA.5 RBD. This suggests that all antibodies induced by the bivalent vaccine – at least with the limit of detection of our assay in polyclonal serum - are in fact cross-reactive. This further suggests that, on a serum antibody level, the bivalent vaccine did not induce a *de novo* response to BA.5.

---

Vaccination against the coronavirus disease 2019 (COVID-19) increased immunity in the population reducing viral transmission and protecting against severe disease. However, continuous emergence of severe acute respiratory syndrome coronavirus 2 (SARS-CoV-2) variants required the implementation of bivalent boosters including the wild-type (WT, D614G) and BA.5 spike. Improved effectiveness of bivalent vs monovalent booster against Omicron sub-variants was reported [1], however limited differences in the immune response are detected [2, 3].

We investigated whether a bivalent COVID-19 booster vaccine comprised of WT + BA.5 spike induced detectable BA.5-specific antibody responses in serum. Samples (n=16) collected before [31 days +/- 63 (0 – 260)] and after [16 +/- 8 days (6 – 31)] receiving the bivalent booster were tested for antibody binding and avidity to the receptor binding domain (RBD) of WT and BA.5. Neutralization of WT and BA.5 viruses was determined. Omicron-specific antibodies were measured by depletion of WT RBD reactive antibodies and assessment of depleted sera against BA.5 RBD.

A significant increase in antibody binding to WT and BA.5 RBD and in neutralization of WT and BA.5 viruses were detected following bivalent booster. There were significant differences in binding of post-booster sera between WT and BA.5 RBDs, however differences in neutralization were not significant. Pre- and post-booster RBD antibody avidity was lower against BA.5 vs WT RBD, which prompted us to look for BA.5 specific antibodies. WT RBD depleted sera lacked reactivity to WT RBD – as expected – and to BA.5 RBD, suggesting that a single exposure to BA.5 antigens by the administration of bivalent vaccine boosters does not elicit robust levels of BA.5 specific serum antibodies.

Reduced sensitivity of antibody tests based on WT viral antigens was detected in naïve individuals after Omicron infection [4]. However, most of the population globally has been infected with ancestral strains and/or exposed to WT antigens through vaccination, hence our results are relevant for the current immune status of the population worldwide. Moreover, our data align with recent results indicating that a monovalent booster with BA.1 vaccine elicits robust spike-specific germinal center B cell responses but low levels of *de novo* B cells targeting variant-specific epitopes [5]. Whether further exposures to Omicron antigens will boost these responses to make them detectable in serum remains to be explored. Importantly, it is likely that cross-reactive antibodies towards Omicron-antigens contribute to protection.

**Figure 1.**
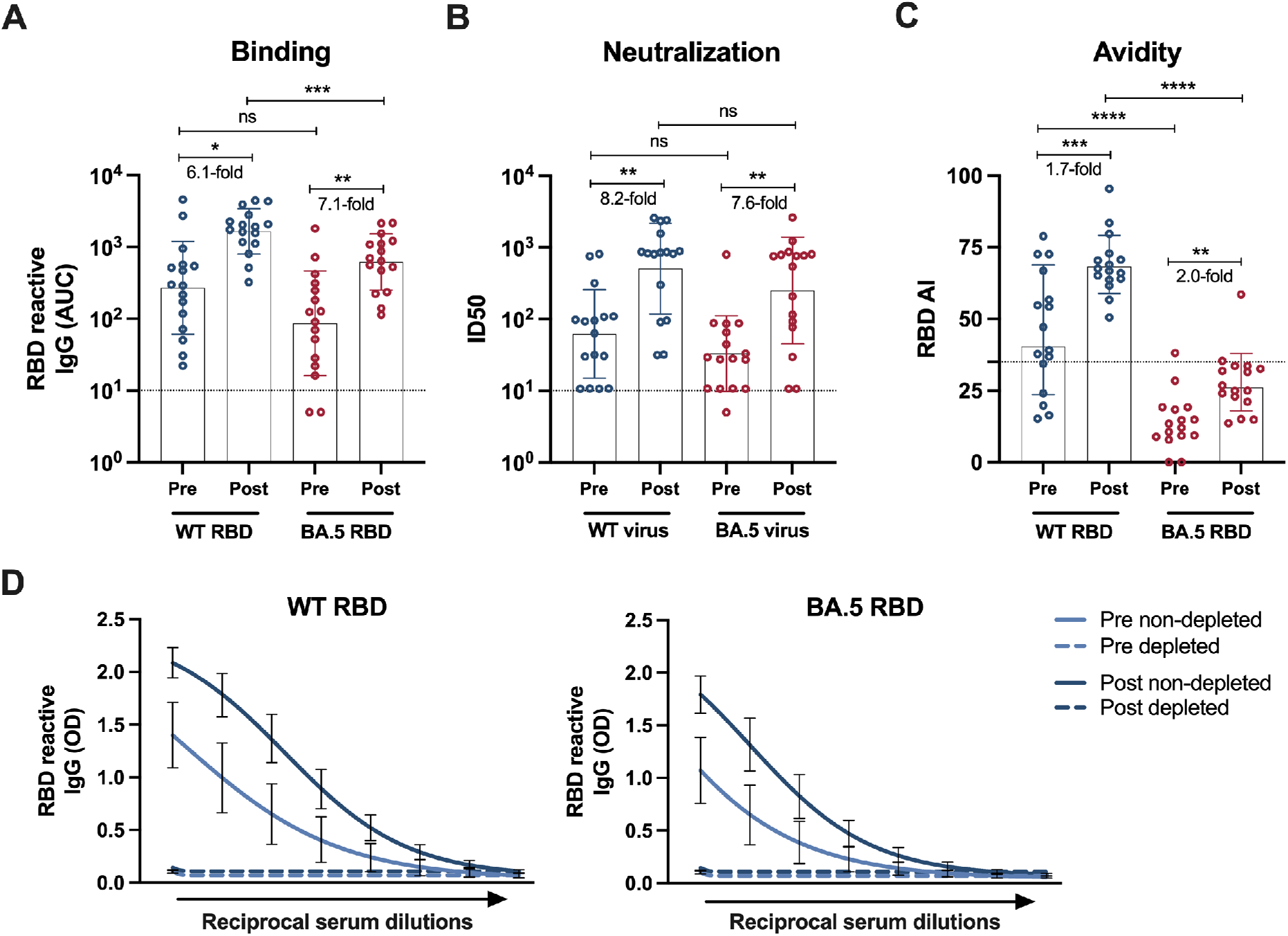
(**A**) Antibody levels expressed as area under the curve (AUC), binding to the recombinant receptor binding domain (RBD) of wild type (WT, blue) and BA.5 (red) SARS-CoV-2, measured in pre- and post-bivalent booster vaccination (WT + BA.5) sera. (**B**) Inhibitory dilution 50% (ID50) of pre- and post-bivalent booster vaccination (WT + BA.5) sera vs WT (blue) and BA.5 (red) live viruses. (**C**) Avidity index (AI) of pre- and post-bivalent booster vaccination (WT + BA.5) sera to WT (blue) and BA.5 RBD (red). (**D**) Reactivity of pre (light blue)- and post (dark blue)- bivalent booster vaccination (WT + BA.5) sera depleted of WT RBD antibodies (dashed lines) or non-depleted (continuous lines) towards WT (left) or BA.5 (right) RBD. Dotted lines in **A-B** indicate the limit of detection of the assay. Dotted line in **C** indicates the threshold for low avidity. In **D**, optical density (OD) in the *y* axis and reciprocal serum dilutions (100-12,800 with 2-fold dilution series) in the *x* axis are shown. A non-linear regression of log (10) transformed data is presented. In **A-C**, average fold change after bivalent vaccine booster is indicated for every pair. A regular one-way analysis of variance (ANOVA) test with Tukey multiple-comparison was performed to compare differences among groups. P values lower than 0.0332 were considered statistically significant with a 95% confidence interval. * P < 0.0332; ** P < 0.0021; *** P < 0.0002; **** P < 0.0001. In **A-D**, geometric mean (GM) plus 95% confidence interval (CI) is shown, n=16.

## Data Availability

All data produced in the present study are available upon reasonable request to the authors

## Supplementary material

### Methods

#### Ethics statement

Sera and data were obtained from two observational studies: the PARIS study (Protection Associated with Rapid Immunity to SARS-CoV-2, approved by the Program for the Protection of Human Subjects at the Icahn School of Medicine at Mount Sinai Institutional Review Board, IRB-20-03374/STUDY-20-00442) and the observational longitudinal clinical sample collection from patients with emerging viral infections research study (approved by the Program for the Protection of Human Subjects at the Icahn School of Medicine at Mount Sinai Institutional Review Board-IRB-17-00791/STUDY-16-01215). Participants from both studies provided informed consent prior to sample and data collection.

#### Study cohort and serum samples

Blood was collected from individuals before and after receiving a Pfizer (n=10) or Moderna (n=6) bivalent vaccine booster of wild type + BA.5 spike. Samples were collected 31 +/- 63 (0 – 260) days before and 16 +/- 8 (6 – 31) days after bivalent booster vaccination. No comorbidities were reported by the participants. Detailed demographic characteristics and vaccination information per individual and per group are indicated in **Supplementary Tables 1 and 2**.

#### Cells and viruses

African green monkey Vero.E6 cells expressing transmembrane protease serine 2 (TMPRSS2) were cultured at 37°C with 5% CO_2_ in Dulbecco’s modified Eagle medium (DMEM, Thermo Fisher Scientific) supplemented with 10% fetal bovine serum (FBS), 1× nonessential amino acids (NEAA), 1% penicillin-streptomycin, 3 μg/ml puromycin (InvivoGen) and 100 μg/ml Normocin (InvivoGen). The SARS-CoV-2 isolate USA-WA1/2020 was used as wild-type reference (BEI Resources, NR-52281) and USA/NY-MSHSPSP-PV58128/2022 provided by the Mount Sinai Pathogen Surveillance Program as representative viral isolate for Omicron BA.5.

#### Recombinant proteins

Recombinant soluble SARS-CoV-2 proteins were expressed using a mammalian cell protein expression system. SARS-CoV-2 spike, RBD, and NP gene sequences from the ancestral strain Wuhan-1 were cloned into a mammalian expression vector pCAGGs. Proteins were produced in Expi293F cells (Thermo Fisher Scientific) using the ExpiFectamine 293 Transfection Kit (Thermo Fisher Scientific) for plasmid transfection. Spike and RBD cell culture supernatants were harvested at 3 days post-transfection, centrifuged at 4000 x g, and filtered. Proteins were purified using Ni^2+^-nitrilotriacetic acid (Ni-NTA) agarose (Qiagen).. Purified proteins were concentrated using Amicon Ultracell centrifugal units (EMD Millipore), and buffer was exchanged to phosphate buffered saline (PBS, pH 7.4). Protein integrity was assessed using sodium dodecyl sulphate (SDS) polyacrylamide gels (5–20% gradient; Bio-Rad). Proteins stocks were stored at -80°C until used.

#### Binding and avidity measurements

Antibody binding and avidity levels were assessed using a research-grade enzyme-linked immunosorbent assay (ELISA). Recombinant receptor binding domain (RBD) from the original Wuhan-Hu-1 SARS-CoV-2 isolate (wild-type, WT) and Omicron (BA.5) SARS-CoV-2 were used. Sera was heat inactivated at 56°C for 1h. Ninety-six-well plates (Immulon 4 HBX; Thermo Scientific) were coated with 50 μl/well of recombinant antigen (2 μg/mL) in phosphate-buffered saline (PBS; pH 7.4, Gibco) and incubated overnight at 4°C. Plates were washed 3 times with 1X PBS supplemented with 0.1% Tween-20 (PBS-T; Fisher Scientific) and blocked with 200 μl/well of 3% non-fat milk PBS-T for 1h at room-temperature (RT). Serum dilutions starting at 1:100 followed by 2-fold dilutions were prepared in 1% non-fat milk PBS-T. Blocking solution was removed and dilutions were added to the plates for 2h at RT. Plates were washed 3 times with PBS-T. For avidity measurements, the same sample was incubated with identical sample dilutions in side-by-side columns; one column was treated with 100 μl/well of 8M Urea while the other was left untreated. Plates were incubated for 10 min, then washed as described above. 50 μl/well of anti-human IgG (Fab-specific, Sigma-Aldrich, A0293) conjugated to horseradish peroxidase (HRP) prepared in 1% non-fat milk PBS-T at 1:3000 dilutions were added for 1h at RT. Plates were washed 3 times and 100 μl/well of o-phenylenediamine dihydrochloride (Sigmafast OPD; Sigma-Aldrich) were added for 10min at RT. To stop the reaction, 50 μl/well of 3 M hydrochloric acid (Thermo Fisher) were added. Optical density (OD) at 490 nm was measured using a Synergy 4 (BioTek) plate reader. Blank wells without serum were used to assess background. Antibody levels expressed as area under the curve (AUC) were calculated by subtracting the background plus three times the standard deviation of the OD. Avidity index (AI) was calculated using the following formula: AI = (Urea-treated sample AUC/Non-treated sample AUC) *100).

#### Antigen coupling to magnetic beads

Carboxyl magnetic beads (RayBiotech, Peachtree Corners, GA) were coated with RBD from WT SARS-CoV-2 at a ratio of 35 μg of antigen per 100 μl of magnetic beads. Mock beads were prepared using PBS (pH 7.4, Gibco) only instead of antigen. Briefly, 1ml of magnetic beads was transferred to a 1.5ml microtube and washed twice with wash buffer consisting of PBS supplemented with 0.1% BSA and 0.05% Tween-20. Beads were resuspended in 1ml of wash buffer and 350 μg of purified N-terminal 6x his tagged WT RBD antigen were added. The mixture was incubated for 2 h at 4°C with constant shaking. Unbound antigen was removed by passage of beads through a magnetic stand. Coupled beads were quenched by the addition of 1ml (1x volume of beads) of 50 mM Tris, (pH 7.4) and incubation at RT with constant shaking. Quenching buffer was removed using the magnetic stand and conjugated beads were washed 4 times with wash buffer. After the final wash, beads were resuspended in 1ml of wash buffer (1x volume of beads) and stored at 4°C prior to use for antibody depletion.

#### Antibody depletion

Depletion of antibodies binding to WT RBD was performed using antigen coupled magnetic beads. Sera were diluted in PBS at a 1:10 ratio and 20 μl of antigen coupled magnetic beads or mock beads were added to 100 ul of pre-diluted sera. The mixture was incubated for 2 hours at 4°C with constant shaking. A magnet stand was used to separate the antibody-antigen-bound beads from the 1x depleted sera. 20 ul of new antigen-coupled beads were added to the 1x depleted sera followed by the same incubation steps as the first round of depletion. 2x depleted sera was used for binding experiments.

#### Microneutralization assay

Serum samples were screened for neutralizing antibodies against ancestral SARS-CoV-2 USA-WA1/2020 and BA.5. Briefly, 96-well plates were seeded with 2×10^4^ Vero.E6 TMPRSS2 cells per well in complete Dulbecco’s modified Eagle medium (cDMEM) 24 hours before infection. Sera were prediluted 1:10 in infection media consisting of minimal essential media (MEM; Gibco, cat. no. 11430-030) supplemented with 2 mM L-glutamine (Gibco, cat. no. 25030081), 0.1% sodium bicarbonate (w/v) (HyClone, cat. no. SH30033.01), 10 mM 4-(2-hydroxyethyl)-1-piperazineethanesulfonic acid (HEPES; Gibco, cat. no. 15630080), 100 U ml−1 penicillin, 100 μg ml−1 streptomycin (Gibco, cat. no. 15140122) and 0.2% bovine serum albumin (BSA) (MP Biomedicals, cat. no. 810063). On the second day, serum dilutions were incubated with 10,000 tissue culture infectious dose 50% (TCID50) of virus per ml for one hour at RT. 120 μl of the virus–serum mix were transfered to every well of Vero.E6 TMPRSS2 plates, followed by incubation for 1h at 37°C. Remdesivir (Medkoo Bioscience inc., cat. no. 329511) was used as a control of virus inhibition at an initial concentration of 10 μg/ml. Viral inoculum was removed and 100 μl/well of the respective serum dilutions plus 100 μl/well of infection media supplemented with 2% fetal bovine serum (FBS; Gibco, Ref. 10082-147) were added. Infection was allowed to proceed for 48 hours at 37 °C. For nucleoprotein (NP) antigen staining, cell monolayers were fixed with 200 μl of 10% paraformaldehyde (PFA) overnight at 4 °C. Cells were washed with 1x PBS then permabilized with 150 μl/well of PBS (pH 7.4) containing 0.1 percent Triton X-100 for 15 minutes at RT. Plates were blocked for 1 hour at RT with 200 μl/well PBS supplemented with 3% BSA. Blocking solution was removed and 100 μl/well of biotinylated monoclonal antibody (mAb) 1C7C7 (mouse anti-SARS NP monoclonal antibody produced at The Icahn School of Medicine at Mount Sinai (ISMMS)) was added at 1 μg/ml in PBS supplemented with 1% BSA for1 hour at RT. Cells were washed with 200 μl/well of PBS twice and 100 μl/well of HRP-conjugated streptavidin (Thermo Fisher Scientific) diluted in PBS supplemented with 1% BSA were added at a 1:2,000 dilution for 1 h at room temperature.Plates were washed twice with PBS and 100 μl/well of Sigmafast OPD were added for 10 minutes at RT. To stop the reaction, 50 μl/well of a 3 M HCl solution (Thermo Fisher Scientific) were added. Optical density (OD, 490 nm) was measured in a Synergy H1 microplate reader (Biotek). For analysis and data representation Prism 9 (GraphPad) was used. Percentage of neutralization, after background subtraction, and compared to the “virus only” control, was calculated. Inhibitory dilution 50% (ID50) was calculated by a non-linear regression curve fit analysis with top and bottom constraints of 100% and 0%, respectively.

#### Statistics

Differences between groups were assessed using a regular one-way analysis of variance (ANOVA) test with Tukey multiple-comparison. P values lower than 0.0332 were considered statistically significant with a 95% confidence interval. Statistical analyses were performed using Prism 9 (GraphPad, USA).

## Financial support

We thank all the participants of the Personalized Virology Initiative’s longitudinal studies for their generous and continued support of research. This effort was supported by the Serological Sciences Network (SeroNet) in part with Federal funds from the National Cancer Institute, National Institutes of Health, under Contract No. 75N91019D00024, Task Order No. 75N91021F00001. The content of this publication does not necessarily reflect the views or policies of the Department of Health and Human Services, nor does mention of trade names, commercial products or organizations imply endorsement by the U.S. Government. This work was also partially funded by the Centers of Excellence for Influenza Research and Surveillance (CEIRS, contract # HHSN272201400008C), the Centers of Excellence for Influenza Research and Response (CEIRR, contract # 75N93021C00014), by the Collaborative Influenza Vaccine Innovation Centers (CIVICs contract # 75N93019C00051) and by institutional funds.

PVI Study Group: Miriam Fried, Sara Morris, Leeba Sullivan, Hala Alshammary, Dalles Andre, Maria C Bermúdez-González, Gianna Cai, Christian Cognigni, Ana Gonzalez-reiche, Hyun Min Kang, Giulio Kleiner, Neko Lyttle, Jacob Mauldin, Brian Monahan, Jessica Nardulli, Annika Oostenink, Jose Polanco, Ashley Salimbangon, Morgan Van Kesteren.

## Author contributions

J.M.C. and F.K. conceptualized study; V.S., C.G., K.S., and the PVI study group enrolled participants, collected data, evaluated surveys, managed IRB approvals, and provided biospecimen and metadata, G.S., T.Y., and J.M.C. performed experiments; H.v.B. performed sequencing of viral isolates; J.M.C., G.S., and A.A. analyzed data; J.M.C., V.S., and F.K. administered the project; F.K., V.S., and H.v.B. provided resources; J.M.C. and F.K. wrote original draft. All authors reviewed, edited and approved the final version of the manuscript, and have had access to the raw data. Members of the PVI study group collected, processed, stored biospecimen, curated metadata, conducted SARS-CoV-2 precision surveillance and assisted with serological antibody measurements.

---

**Supplementary Table 1.** Detailed description of samples used and demographics per individual.

| Participant ID<sup>1</sup> | Age Bracket | Sex | Ancestry | Time points included in this study | SARS CoV-2 infection prior to vaccination | Primary Vaccine type | Bivalent booster type |
| --- | --- | --- | --- | --- | --- | --- | --- |
| BVB-1 | 35-39 | Male | Caucasian<br>Not Hispanic or Latino | Pre-Boost,<br>Post Boost | No | Pfizer | Moderna |
| BVB-2 | 55-59 | Male | Caucasian<br>Hispanic or Latino | Pre-Boost,<br>Post Boost | Yes | Moderna | Pfizer |
| BVB-4 | 45-49 | Male | Caucasian<br>Not Hispanic or Latino | Pre-Boost,<br>Post Boost | Yes | Pfizer | Pfizer |
| BVB-5 | 40-44 | Female | Other<br>Not Hispanic or Latino | Pre-Boost,<br>Post Boost | Yes | Pfizer | Moderna |
| BVB-6 | 45-49 | Male | Other<br>Not Hispanic or Latino | Pre-Boost,<br>Post Boost | Yes | Pfizer | Moderna |
| BVB-7 | 35-39 | Female | Caucasian<br>Not Hispanic or Latino | Pre-Boost,<br>Post Boost | Yes | Pfizer | Pfizer |
| BVB-8 | 40-44 | Female | Caucasian<br>Not Hispanic or Latino | Pre-Boost,<br>Post Boost | Yes | Pfizer | Pfizer |
| BVB-9 | 65-69 | Male | Caucasian<br>Not Hispanic or Latino | Pre-Boost,<br>Post Boost | Yes | Pfizer | Pfizer |
| BVB-10 | 40-44 | Female | Caucasian<br>Not Hispanic or Latino | Pre-Boost,<br>Post Boost | No | Pfizer | Moderna |
| BVB-11 | 30-34 | Female | Caucasian<br>Not Hispanic or Latino | Pre-Boost,<br>Post Boost | Yes | Pfizer | Pfizer |
| BVB-12 | 60-64 | Female | Caucasian<br>Not Hispanic or Latino | Pre-Boost,<br>Post Boost | Yes | Pfizer | Pfizer |
| BVB-13 | 40-44 | Female | Caucasian<br>Not Hispanic or Latino | Pre-Boost,<br>Post Boost | No | Pfizer | Pfizer |
| BVB-14 | 70-79 | Female | Caucasian<br>Not Hispanic or Latino | Pre-Boost,<br>Post Boost | No | Pfizer | Moderna |
| BVB-15 | 40-44 | Female | Caucasian<br>Not Hispanic or Latino | Pre-Boost,<br>Post Boost | Yes | Pfizer | Moderna |
| BVB-16 | 45-49 | Female | Caucasian<br>Not Hispanic or Latino | Pre-Boost,<br>Post Boost | No | Pfizer | Pfizer |
| BVB-17 | 70-79 | Male | Caucasian<br>Hispanic or Latino | Pre-Boost,<br>Post Boost | Yes | Pfizer | Pfizer |

Summary of the demographic, infection and vaccination histories of participants from whom serum samples were analyzed. The following abbreviations are used in the table, Pre-Boost: serum collected before the bivalent vaccine booster, Post Boost: serum collected after the bivalent vaccine booster.
<sup>1</sup>This is a code (bivalent booster/BVB abbreviation plus random number), not a patient ID.

**Supplementary Table 2.** Age and sex characteristics of individuals per group.

|  |  |  |
| --- | --- | --- |
| Overall | Number of participants | 16 |
|  | Average age (min – max) | 48 (32 – 73) |
|  | Average days before bivalent booster +/- stdv (min – max) | 31 +/- 63 (0 – 260) |
|  | Average days after bivalent booster +/- stdv (min – max) | 16 +/- 8 (6 – 31) |
| Male | Number of participants | 6 |
|  | Average age (min – max) | 53 (37 – 73) |
|  | Average days before bivalent booster +/- stdv (min – max) | 13 +/- 17 (0 – 38) |
|  | Average days after bivalent booster +/- stdv (min – max) | 17 +/- 8 (10 – 28) |
| Female | Number of participants | 10 |
|  | Average age (min – max) | 45 (32 – 73) |
|  | Average days before bivalent booster +/- stdv (min – max) | 41 +/- 78 (2 – 260) |
|  | Average days after bivalent booster +/- stdv (min – max) | 15 +/- 9 (6 – 31) |

The following abbreviations are used in the table: standard deviation (stdv), minimum (min), maximum (max).

